# Acute Exercise-Induced Changes in Gut Microbiome Composition, Function, and Gut-Derived Stool and Plasma Metabolome Across Obesity Phenotypes in Young-Adult Women: A Pilot Study Protocol

**DOI:** 10.64898/2026.07.10.26357758

**Authors:** Carmen P. Ortega-Santos, Daniel Kerchner, Ali Rahnavard, Keith A. Crandall

**Affiliations:** Exercise and Nutrition Sciences Department, Milken Institute School of Public Health, Washington, DC, USA; Computational Biology Institute, Department of Biostatistics and Bioinformatics, Milken Institute School of Public Health, The George Washington University, Washington, DC, USA

**Keywords:** Gut Microbiome, Metabolome, Exercise, Women, Obesity, Pilot Projects, Clinical Trial Protocol

## Abstract

**Background:** Almost 1 in 2 adults in the US has obesity, with women facing the highest prevalence of severe obesity. Emerging evidence shows that the gut microbiome and its metabolites play a key role in metabolic regulation, acting as signals in active metabolic tissues such as adipose and skeletal muscle, thereby contributing to the deterioration of cardiometabolic health in individuals with obesity. Recent findings suggest that functional characteristics of the gut microbiome, rather than compositional changes alone, may partially explain why some individuals experience greater metabolic benefits from exercise than others. We designed a pilot trial to assess the acute response to an exercise bout across distinct obesity phenotypes and to identify microbial signatures associated with lifestyle interventions.

**Methods:** We proposed a pilot trial in which 40 young adults (21 to 40 years old) with distinct exercise (< 150 minutes per week or ≥ 4 hours per week) and body compositions (body mass index [BMI] 18.5 to 24.99 or ≥ 30 kg/m^2^) would undergo an acute exercise bout. The outcomes of this pilot trial are as follows: (1) Examine the effects of a 30-minute bout of moderate-intensity aerobic exercise (60–70% heart rate reserve) on the abundance and functional activity of short-chain fatty acid (SCFA)-producing gut bacteria across different obesity phenotypes in women; (2) Assess the acute effects of the same exercise bout on SCFA concentrations in stool and circulating plasma metabolomic profiles. Discussion: The results of this pilot study will inform the feasibility of a larger trial to establish the gut-synthesized metabolites of exercise response across obesity heterogeneity in young adult women.

**Trial Registration:** This study was registered on October 16^th,^ 2024, at clinicaltrials.gov.gov, ID: NCT06691100

## Introduction

### Background

Obesity is a chronic, heterogeneous disease, and understanding its biological underpinnings is essential for developing more effective, personalized, and sustainable strategies to prevent, treat, and improve obesity and cardiometabolic diseases. Almost 1 in 2 adults in the US has obesity, especially women, who face the highest prevalence of severe obesity.^1,2^ Obesity increases the risk of chronic diseases like cardiovascular disease, diabetes, and cancer, contributing over $200 billion to healthcare costs annually in the US.^3^ Emerging evidence shows that the gut microbiome and its metabolites play a key role in metabolic regulation, acting as signals in active metabolic tissues such as adipose and skeletal muscle. Adults with obesity exhibit reduced microbial diversity and richness, a lower healthy-to-pathogenic bacterial ratio (i.e., dysbiosis), and disruptions in metabolite synthesis, leading to lower fecal and plasma short-chain fatty acid (SCFA) abundances and elevated plasma branched-chain amino acid levels.^4–8^

Disruption of the gut microbiome composition is associated with the development of obesity, insulin resistance, and type 2 diabetes mellitus (T2DM).^9–15^ The underlying biological mechanisms have yet to be defined, but a greater synthesis of gut metabolites, such as short-chain fatty acids (SCFA), is correlated with metabolic benefits, including improved insulin sensitivity and glucose metabolism.^6,7,16–24^ Until recently, most studies examining the effects of lifestyle factors on gut microbiome composition focused on diet; however, evidence indicates that as little as 4 weeks of aerobic exercise can significantly increase the relative abundance of SCFA-producing bacteria in individuals with obesity, independent of dietary intake.^25–30^ Yet the impact of aerobic exercise on gut microbiome-related metabolites and the extent to which these exercise-induced metabolites improve adiposity and insulin sensitivity in adults with obesity remain poorly studied. This knowledge gap underscores the importance of examining this pathway closely to better inform more personalized approaches to treating obesity.

The beneficial multiorgan effects of aerobic exercise on metabolic homeostasis are well-defined across health and obesity.^31–33^ Most prior studies on the effects of exercise on the gut microbiome are cross-sectional and examine healthy-weight professional and recreational athletes, among whom the gut microbiome is diverse and abundant in SCFA-producing bacteria (*Roseburia spp* and *Ruminococcus spp*).^34,35^ More recently, gut microbiome composition similar to that of healthy and athletic populations has been observed following at least four weeks of structured aerobic exercise training in sedentary individuals.^25,27–30,36,37^ While these studies make important strides in our understanding of exercise-induced changes in gut microbiome composition and gut-synthesized metabolites, many overlook key confounding influences (namely, diet), complicating the isolation of the effects of exercise *per se*. Also, while demonstrating that aerobic exercise positively influences gut microbiome composition in obesity, prior studies often neglect to assess whether these microbial changes enhance cardiometabolic health. Most importantly, no studies have analyzed the integration of shotgun metagenomics and gut metabolites to evaluate gut microbiome functional activity or to monitor fluctuations during exercise training, thereby hindering our understanding of how these changes relate to individual response variability. This variability in responses reflects the broader heterogeneity within obesity, in which individuals with similar body mass index (BMI) exhibit vastly different cardiometabolic profiles, microbiome signatures, and responses to interventions.^25,38,39^ Recent findings suggest that functional characteristics of the gut microbiome, rather than compositional changes alone, may partially explain why some individuals experience greater metabolic benefits from exercise than others.^9,25^ This highlights the need to move beyond composition to examine microbial functionality in exercise. Thus, incorporating integrative omics analyses to understand individual variability in lifestyle intervention responses will address gaps and identify gut-derived metabolites as potential biomarkers for obesity exercise prescription, refining personalized interventions.^9,40,41^ We plan to examine the effects of acute aerobic exercise on gut microbiome composition, functional activity, and gut-derived metabolites, and their links to improved metabolic regulation in sedentary young adult women with obesity. Findings will lay the groundwork for identifying novel therapeutic targets in the gut microbiome and its derived metabolome to improve treatment and prognosis of cardiometabolic disease among adults with obesity. This work can help develop personalized lifestyle interventions for obesity heterogeneity, moving beyond a *one-size-fits-all* approach to improve cardiometabolic health.

Acute exercise has been proposed as a physiological stress test that might serve as a valuable experimental model for better understanding interindividual variability in the gut microbiome response to exercise interventions.^42^ Unlike long-term training studies,^43^ which reflect cumulative adaptations influenced by changes in body composition, diet, and behavior, acute exercise allows investigators to isolate the immediate physiological perturbation induced by exercise itself. Grosicki et al. propose that these short-term responses may provide insight into how transient shifts in gut motility, intestinal perfusion, metabolite production, immune signaling, and gut barrier function interact with the microbiome before chronic adaptation occurs.^42^ Importantly, examining the response to a single exercise bout may help identify early-response biomarkers that distinguish “responders” from “non-responders” to longer-term exercise interventions.^9^ Such biomarkers could include exercise-induced changes in microbial metabolites (e.g., short-chain fatty acids), inflammatory markers, or taxa-specific abundance patterns. This approach may be especially informative in populations with obesity, where baseline microbiome composition and metabolic dysfunction may alter responsiveness, and in women, who remain underrepresented in current exercise–microbiome literature despite potential sex-specific physiological responses.^37,44^ Ultimately, acute exercise challenge models may provide a mechanistic framework for more accurately predicting individualized responses to exercise lifestyle interventions and for supporting the development of more personalized exercise prescriptions.

### Objectives

This pilot study aims to evaluate the acute effects of a standardized bout of aerobic exercise on gut microbiome composition and metabolomic responses in adult women stratified by adiposity phenotype.

Specifically, the study aims to:

1. Examine the effects of a 30-minute bout of moderate-intensity aerobic exercise (60–70% heart rate reserve) on the abundance and functional activity of SCFA-producing gut bacteria across different obesity phenotypes in women.
2. Assess the acute effects of the same exercise bout on SCFA concentrations in stool and circulating plasma metabolomic profiles.

### Trial Design

This study is a single-arm intervention clinical pilot trial with predefined phenotypic group stratification. All participants complete the same standardized acute exercise protocol, and between-phenotype differences in physiological and molecular responses are assessed as exploratory analyses. The study population consists of adult premenopausal women stratified by habitual physical activity level (sedentary vs. exercisers) and obesity phenotype (lean [BMI 18.5-24.99] vs. with obesity [BMI ≥ 30]).

The study includes three laboratory visits. Visit 1 includes informed consent, baseline assessments of body composition, demographics, muscular strength, and cardiorespiratory fitness. Participants are instructed in procedures and provided with materials for stool sample collection, food intake tracking, and physical activity monitoring.

Between Visit 1 and Visit 2 (5–10 days), participants collect a baseline stool sample at home at least 48 hours after Visit 1 and any time before Visit 2. During this period, physical activity is monitored using accelerometry, and dietary intake is recorded using electronic food logs.

Visit 2 consists of a standardized acute exercise session performed in a controlled laboratory environment. Blood samples are collected immediately before exercise and within 3 minutes after completion of exercise to assess acute metabolic responses.

Following Visit 2, participants collect a second stool sample at home during the 24-hour post-exercise period while continuing physical activity and dietary monitoring. Participants return to the laboratory at 24 hours post-exercise (Visit 3), where the stool sample is submitted, and a final blood sample is obtained to assess delayed circulating gut-derived metabolomic responses.

This study is designed to generate preliminary mechanistic evidence linking acute exercise to gut microbiome–metabolome interactions across obesity phenotypes.

## Participants, Interventions and Outcomes

### Study Setting

This study will be conducted in the Washington, DC, USA, metropolitan area at the Milken Institute School of Public Health at The George Washington University. All study procedures, including participant recruitment, acute exercise testing, biological sample collection, processing, and analyses, will be performed within the university’s exercise physiology and translational research facilities.

### Participant Eligibility

Participants in this study will include individuals assigned female sex, aged 21 to 40 years, and we will include women with a body mass index between 18.5 and 24.99 kg/m^2^ or with a BMI ≥ 30 kg/m^2^. Participants will be stratified into four phenotypic groups based on habitual physical activity level and obesity phenotype: Lean-exerciser (Ln-EX), lean-sedentary (Ln-SED), obesity-exerciser (Ob-EX), and obesity-sedentary (Ob-SED). Physical activity classification will be based on self-report, with sedentary individuals defined as those who engage in <150 minutes of moderate-intensity physical activity per week or <75 minutes of vigorous-intensity physical activity per week. Physically active individuals (i.e., exercisers) will be defined as engaging in ≥4 structured exercise sessions per week (≥ 240 minutes/week) of endurance (e.g., running, cycling, triathlon), strength (e.g., powerlifting), or team sports (e.g., soccer, rugby).

Exclusion criteria include pregnancy, lactation, or ≤2 years postpartum; use of dietary supplements with known gut microbiome-modulating effects (e.g., probiotics, prebiotics, fiber supplements) within 1 month prior to enrollment; antibiotic use (oral or vaginal) within 1 month prior to enrollment; and presence of chronic diseases known to affect gut microbiome composition, including endocrine disorders (e.g., polycystic ovarian syndrome), gastrointestinal disease, cancer, or cardiometabolic disease (e.g., type 2 diabetes, hypercholesterolemia). Participants using medically prescribed chronic medications will be excluded.

### Recruitment and Consent

Participants will be recruited from the Washington, DC metropolitan area using community-based and institutional strategies, including outreach through local fitness facilities, sports organizations, student groups, and weight-management clinics. Additional recruitment will be conducted through social media advertising, university newsletters, community listservs, and online platforms. Clinical referrals will also be obtained through collaboration with a lifestyle medicine physician at the George Washington University Medical Faculty Associates.

Interested individuals will complete an online prescreening questionnaire to assess preliminary eligibility. Individuals meeting initial screening criteria will be contacted by study personnel via email, text, or phone.

Eligible participants will be provided with study information prior to their first visit. Written informed consent will be obtained in person at the first laboratory visit by trained research staff in a private setting. Participants will be informed of study procedures, risks, and benefits, and the voluntary nature of participation. Participants will provide informed consent for the collection, storage, and biobanking of biological samples (blood and stool), as well as for their use in future ancillary and secondary analyses related to gut microbiome and metabolomic research. All samples will be de-identified and stored in accordance with institutional procedures and regulatory requirements.

### Intervention

All participants will complete a single 30-minute bout of acute aerobic exercise on a clinical cycle ergometer (Monark 939E, Sweden). Exercise intensity will be prescribed at a target heart rate (*Target HR*) with an intensity (*I*) of 60–70% of heart rate reserve (HRR), calculated individually using the Karvonen method,^45^ based on resting (*HR_rest_*) and maximal (*HR_max_*) heart rates obtained during the baseline cardiopulmonary fitness assessment:

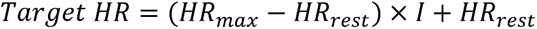

Each session will include a standardized 5-minute low-intensity warm-up followed by the 30-minute exercise bout. All exercise sessions will be conducted in a controlled laboratory environment with standardized conditions, including the absence of music or external auditory stimulation. Heart rate will be continuously monitored to ensure adherence to the prescribed intensity.

Participants will be instructed to refrain from strenuous physical activity and alcohol consumption for 24 hours prior to each study visit and to complete the exercise session following a minimum 5-hour fasting period. Approximately 500 mL of water will be provided prior to exercise to maintain hydration.

### Outcomes

This study will assess outcomes to determine changes in gut microbiome composition, function, and metabolome in response to an acute aerobic exercise bout. The primary outcomes for this study are as follows:

1. Fecal microbiome composition (taxonomic profiling)
2. Relative abundance of short-chain fatty acid (SCFA)-producing bacterial taxa
3. Functional potential of the fecal microbiome, including biosynthetic gene cluster (BGC) activity
4. Stool concentrations of SCFA
5. Plasma concentrations of SCFA

Secondary outcomes for this study:

1. *Dietary intake:* Dietary intake will be assessed throughout the study using repeated self-administered dietary records completed via the free, app-based Cronometer platform. Participants will be instructed to record all food and beverage intake throughout the study. Dietary intake data will be used to characterize habitual diet and to control for potential confounding effects in analyses of gut microbiome and metabolomic outcomes.
2. *Physical Activity:* Physical activity will be assessed using both self-reported and objective measures. At baseline, participants will report habitual physical activity levels. Objective physical activity will be continuously monitored throughout the study using wearable devices to quantify the intensity, duration, and frequency of movement, for exploratory and confounding-control analyses.
3. *Cardiovascular fitness level:* Cardiovascular fitness will be assessed at baseline using a graded exercise test performed on a cycle ergometer to determine maximal oxygen uptake (VO₂max). VO_2max_ will be used (1) to individualize exercise intensity prescription for the experimental trial based on HRR, and (2) to classify participants’ cardiovascular fitness level for exploratory phenotypic analyses.
4. *Muscular Strength Assessment:* Upper- and lower-body muscular strength will be assessed at baseline using handgrip dynamometry and an isometric mid-thigh pull test. These measures will be used to characterize baseline musculoskeletal fitness and classify participants into relative-strength fitness strata for exploratory phenotypic analyses.
5. *Fasting Blood Glucose:* Fasting blood glucose will be measured using a point-of-care analyzer. Glucose levels will be used to characterize participants’ baseline glycemic status and metabolic phenotype.
6. *Fasting Blood Lipids:* Fasting blood lipid profiles will be assessed using a point-of-care lipid analyzer. Measurements will include total cholesterol, high-density lipoprotein (HDL), and triglycerides. These measures will be used to post-hoc characterize participants’ cardiometabolic status and phenotype description.
7. *Body Composition:* Body composition measures will be assessed via dual-energy X-ray absorptiometry (DXA), including total body fat percentage, fat mass, lean mass, and regional adiposity indices. These data will characterize adiposity phenotype and support exploratory analyses.
8. *Sleep quality:* Sleep quality will be assessed using the Pittsburgh Sleep Quality Index (PSQI), to characterize habitual sleep patterns and quality.
9. *Weight and Lifestyle Inventory (WALI):* Retrospective weight history and weight management patterns will be assessed using the WALI questionnaire to characterize obesity chronicity, prior lifestyle behaviors, and phenotype.

### Participant Timeline

The schedule of enrolment, interventions, and assessments is presented in Table 1. The study includes three laboratory visits and at-home sample-collection periods, during which stool samples are collected, physical activity and dietary intake are monitored. Biological sampling and exercise testing occur at standardized time points relative to the acute exercise bout.

**Table 1.** Schedule of Study Procedures by Visit.

| Table 1. Schedule of Study Procedures by Visit |  |  |  |  |  |
| --- | --- | --- | --- | --- | --- |
| Study Phase | Screening | Baseline (Visit 1) | Between visits | Exercise Intervention (Visit 2) | 24h follow up (Visit 3) |
| Eligibility | X |  |  |  |  |
| Consent |  | X |  |  |  |
| Body Composition |  | X |  |  |  |
| Fitness testing |  | X |  |  |  |
| Stool sample |  |  | X |  | X |
| Blood sample |  |  |  | X | X |
| Exercise intervention |  |  |  | X |  |
| Activity (diet and physical activity) monitoring |  | X | X | X | X |

### Sample Size

FITGut-W is a pilot, proof-of-concept study and is not powered to test formal hypotheses. The primary aim is to generate preliminary estimates of effect sizes and variability in microbiome responses to acute exercise.

Data from this study will be used to estimate within-subject and between-phenotype effect sizes (e.g., Cohen’s *d* or standardized mean differences). These estimates will inform sample size calculations for a future fully powered trial.

## Data Collection, Management, and Analysis

### Data Collection Methods

#### Biological Samples

##### Blood collection

Approximately 7–10 mL of blood will be collected at three time points: baseline (pre-exercise), 3 minutes post-exercise, and 24 hours post-exercise. Samples will be collected into EDTA tubes (Fisher Scientific, USA) by a trained phlebotomist using a 21-gauge butterfly needle under standard aseptic procedures.

Following collection, blood samples will be placed on ice and processed within 30–45 minutes. Samples will be centrifuged to separate plasma, which will then be aliquoted and stored at –80 °C until subsequent mass spectrometry analysis for metabolite identification. SCFAs will be quantified using targeted metabolomics approaches. In addition, biospecimens will be stored for future untargeted metabolomics analyses to enable exploratory discovery of circulating and stool-derived metabolites associated with acute exercise responses.

##### Stool collection

A stool sample will be collected using an at-home stool collection kit (Fisher Scientific, MA, USA). Participants will use a DNA/RNA-stabilizing collection tube (Zymo Research, CA, USA) to store stool for metagenomic analysis. The sample will be stored in the participants’ home freezers until delivery. The baseline stool sample will be collected at least 48 hours after the baseline visit to avoid any potential acute effects of the baseline exercise tests on the gut microbiome, and before visit 2. The second stool sample will be collected within 24 hours after visit 2. The samples collected in the DNA/RNA stabilizing collection tube will be stored at 4°C until processing for DNA extraction. The entire stool sample will be processed in a biosafety cabinet, aliquoted into 50 mL and 2 mL tubes, and stored at –80°C until further analysis. We will measure the pH of the stool sample and classify it using the Bristol scale.

Initial processing involves extracting genomic DNA from the collected stool samples using ZymoBIOMICS DNA Microprep Kits (Zymo Research, CA, USA), thereby producing high-quality DNA suitable for downstream sequencing. We will quantify the amount and quality of extracted DNA using the Qubit 4 Fluorometer (Thermo Fisher Scientific, MA, USA) and the Bioanalyzer (Agilent, CA, USA) following the manufacturer’s instructions. Libraries will be prepared using the Nextera XT kit and sequenced using shotgun metagenomics sequencing with the NextSeq 2000 High-Output 300-cycle kits (Illumina), along with the ZymoBIOMICS Gut Microbiome Standard (Zymo, CA, USA), for benchmarking and validation of the NGS microbiome workflow. For quality control, whole-metagenomic shotgun sequencing reads from these samples will be filtered and trimmed to remove low-quality reads using FASTQC/MultiQC^46^ and Trimmomatic.^47^ Then, quality-filtered reads will be mapped to the hg38 human genome reference assembly to partition host (human) reads from the remaining reads, with a target of ∼2-4 million microbial reads per sample after QC. The remaining reads will be classified using PathoScope2,^48^ Kraken2,^49^ and/or seq.^50^ The resulting species will have their functional activity determined using bgcLens, which identifies BGCs through a genomic language model (seqSight) trained on BGC databases, including MIBiG.^51^ We will calculate the relative abundance of SCFA-producing bacterial species by aggregating the abundances of the known SCFA producers (e.g., *Roseburia spp., Lactobacillus spp., Ruminococcus spp.*) and dividing by them the total abundance of all species in each sample.

For metabolomics analysis, stool samples will be dried in an evaporator. We will collect 25-30 mg of dry fecal samples and add 200 µL of extraction buffer (methanol/water 50/50) containing 200 ng/mL of debrisoquine (DBQ) as an internal standard for positive mode and 200 ng/mL of 4-nitrobenzoic acid as an internal standard for negative mode. Metabolites in serum samples will be extracted with methanol and centrifuged. Homogenates will be subjected to automated biochemical extraction and analysis by gas chromatography and high-resolution tandem mass spectrometry (MS/MS). Raw data will be extracted, peaks will be identified, and processed using massSight.^52^ Metabolites will be identified by comparison with library entries for purified standards or recurrent unknown entities. We will use profiling techniques to measure a comprehensive range of metabolites (e.g., free fatty acids, bile acids, purines, and central metabolites), both annotated and unannotated. Three types of controls will be included: a pool of small portions of each experimental sample, serving as a technical replicate throughout the platform run; extracted water samples (process blanks); and a mixture of standards spiked into every analyzed sample, allowing for monitoring of instrument performance and drifts in metabolite descriptors (m/z and retention time). We perform error correction, normalization, drift correction, and quality control on the metabolite data, including data cleaning and curation, to enhance interpretation. Clinical and metabolomics data will be analyzed to identify metabolites of interest and key clinical features associated with significant changes in metabolomics between the treatment groups. Aliquots will be preserved for future analysis, contingent upon funding availability, to investigate additional metabolites of interest, including tryptophan, secondary bile acids, and branched-chain amino acids, all associated with GM activity.

#### Clinical and Physiological Tests

##### Cardiovascular fitness test (VO_2max_)

All participants will complete the cardiovascular fitness test in a fasted state (≥ 8 hours) during the final component of the baseline visit. Maximal oxygen uptake (VO₂max) will be assessed using a graded exercise test performed on an electronically braked cycle ergometer (Monark 939E, Sweden) with continuous respiratory gas exchange measurements obtained via a metabolic cart (COSMED, Italy). The protocol will include 2 minutes of resting gas-exchange measurements, followed by a 5-minute warm-up at 20 watts. The test will then proceed with incremental increases of 15 watts per minute, starting at 20 watts, maintaining 50 rpm at all times, until volitional exhaustion. VO₂max will be determined based on the identification of VO₂max, defined as the highest 15-second averaged oxygen uptake values achieved during the test. Attainment of VO₂max will be supported by standard physiological criteria, including a respiratory exchange ratio (RER) ≥ 1.1, attainment of a plateau in heart rate or maximal heart rate, or a plateau in oxygen uptake despite increasing workload.

##### Body composition

All participants will be administered a total body composition assessment using basic anthropometric measurements and a Dual-Energy X-ray Absorptiometry (DXA; GE Lunar iDXA) in a fasted state (≥ 8 hours) during the baseline visit. Participants’ weight will be measured to 0.1 kg with a calibrated scale in light clothing and no shoes. Height will be measured to 0.1 cm with a Seca stadiometer, with participants barefoot and standing. Abdominal circumference will be measured at the umbilicus or midway between the lowest rib and iliac crest with a non-stretchable tape. Hip circumference, at the widest part, will be measured to 0.1 cm. If measurements differ by ≥0.5 cm, a third measurement will be taken, and the average of the three measurements will be used. Following anthropometric measurements, participants lie supine while the scanner measures bone mineral density, lean tissue, and fat mass. DXA scans provide detailed regional and total body composition data, including fat mass (kg, %), free fat mass (kg), and visceral area (cm^2^).

##### Strength tests

Participants will complete two strength assessments at baseline to evaluate upper- and lower-body maximal force production. Handgrip strength will be assessed using a handheld dynamometer. Participants will perform three maximal trials with their dominant hand, with at least 1 minute of rest between trials. The highest value recorded will be used for analysis. Lower-body strength will be assessed using an isometric mid-thigh pull performed on force plates (Hawkin Dynamics, Maine, USA). Participants will first complete a familiarization trial at approximately 50% of perceived maximal effort. For testing, participants will be positioned with the bar fixed at mid-thigh height to ensure consistent posture across trials. Participants will then perform three maximal isometric pulls, with standardized instructions and sufficient rest between trials. Peak force will be recorded using the force plate system software, and the highest value will be used for analysis.

#### Behavioral Measures

##### Diet

Participants will be instructed to record all food and beverage intake throughout the study, beginning the day following the baseline visit. Dietary intake will be captured using the Cronometer web- and smartphone-based application, for which participants will be provided with an individual account at no cost. Members of the research team will have access to participants’ dietary records to monitor compliance and ensure completeness of data collection throughout the study period. Upon study completion, dietary data will be downloaded from Cronometer and manually entered into the Nutrient Data System for Research (NDSR) to analyze energy intake, macronutrient composition, fiber intake, and selected micronutrients. The Healthy Eating Index^53,54^ will also be calculated for descriptive analysis. Dietary intake data will be used to characterize habitual diet and to account for potential confounding effects in analyses of gut microbiome and metabolomic outcomes.

##### Physical activity

Physical activity will be assessed using both self-reported and objective measures. Participants will report habitual physical activity, including minutes of moderate and vigorous activity and participation in structured sports or exercise, as part of baseline characterization. Objective physical activity will be continuously monitored throughout the study using an ActiGraph accelerometer (wGT3X-BT, Ametris, Pensacola, FL) worn on the right hip. Participants will be instructed to wear the device at all times, except during sleep and water-based activities (e.g., showering or swimming). Accelerometer data will be used for post-hoc quantification of physical activity levels, including intensity, duration, and frequency of movement.

#### Questionnaires

Standardized questionnaires will be administered at baseline to characterize sleep, weight history, and participant demographics. Sleep quality will be assessed using the Pittsburgh Sleep Quality Index (PSQI),^55^ a validated questionnaire that evaluates habitual sleep patterns and sleep quality over the previous month. Weight history and lifestyle patterns will be assessed using the Weight and Lifestyle Inventory (WALI)^56^ to characterize obesity trajectory and weight management behaviors. In addition, participants will provide demographic information, including age and relevant health history. Menstrual cycle status will also be assessed at baseline, including the date of the last menstrual cycle and cycle duration.

### Statistical Analysis

We will perform descriptive statistics using the demographics and medical history data. For all continuous measures, we will test for normality using the Kolmogorov-Smirnov test and q-q plots.

We will also conduct per-protocol analyses, removing those individuals who were not adherent to the intervention protocol. All microbiome and metabolome data will be centered log-ratio (CLR) transformed and appropriately log-transformed to address the compositional nature of omics data. Multi-omics data are characterized by zero-inflation and high dimensionality. We will utilize machine learning-based analysis tools designed for multi-omics data including: 1) *omeClust*,^57^ an -omics community detection tool that uses multi-resolution clustering to identify patterns in the omics dataset and explanatory variables; 2) *Tweedieverse*,^58^ a tool which implements a statistical framework based on the Tweedie distribution to perform differential analysis of multi-omics to identify associations between omics features and key outcomes of interest (e.g., body composition); 3) *omePath*,^59^ software that performs omics pathway enrichment analysis to investigate enriched or depleted functional pathways (or groups of features), and 4) *btest,*^59^ *a* tool to link, rank, and visualize associations among -omics features. Curated metabolite intensities generated by liquid chromatography-mass spectrometry (LC-MS) and participant features will be used as inputs to *omeClust*.

We will compare within-group (pre- vs. post-intervention) and between-group differences using a modified general linear model implemented in Microbiome Multivariable Associations with Linear Models 2 (MaAsLin2).^60^ The main fixed-effect will be the group × time interaction, with group and time (pre vs. post) also included as fixed effects to model main and interaction effects. We will include participant ID as a random effect to account for within-subject correlation due to repeated measures. The dietary fiber intake covariate will be included as an additional fixed effect to adjust for potential confounding. We will compare within-group (pre- vs. post-intervention) and between-group differences in SCFA in stool and plasma using linear mixed-effects models. The fixed effects will include group, time (pre vs. post), and the group × time interaction term, with participant ID included as a random effect to account for repeated measures. All nominal p-values will be corrected for multiple hypothesis testing using Benjamini-Hochberg false discovery rate (FDR) procedures, with significance set at q□<□0.05.

### Data Management

All study data will be collected and managed using a secure, password-protected electronic data capture system REDCap.^61^ Hard copies of signed informed consent forms will be stored in a locked filing cabinet in a secure, access-restricted research facility. Access will be limited to authorized study personnel to maintain participant confidentiality. Participant data used for analysis will be assigned unique study IDs, and no direct identifiers will be stored with research data. The key linking participants to study IDs will be maintained through a secure, restricted-access key file.

Data from questionnaires, clinical assessments, and laboratory procedures will be entered directly into the electronic database or transferred from source documents by trained research personnel. Data entry will be subject to built-in validation checks and periodic quality control reviews to ensure accuracy and completeness.

Biological samples (blood and stool) will be labeled using coded identifiers and stored in accordance with institutional biobanking procedures to maintain traceability while preserving participant anonymity. Linkage between biological specimens and clinical data will be maintained through a secure, restricted-access key file.

All study data will be stored on secure institutional servers with access limited to authorized study personnel. Data will be retained in accordance with institutional and regulatory requirements.

## Ethics and Dissemination

### Research Ethics Approval

Ethical approval for this study has been obtained from The George Washington University Institutional Review Board, NCR 245983.

### Protocol Amendments

Any modifications to the study protocol will be submitted to the George Washington University Institutional Review Board (IRB) for review and approval prior to implementation. Approved amendments will be incorporated into the protocol and documented accordingly.

If protocol modifications affect participants who have already been enrolled, re-consent will be obtained as appropriate before continued participation. Relevant protocol updates will also be reflected in the ClinicalTrials.gov registry.

### Confidentiality

All participant information will be kept confidential and managed in accordance with institutional and regulatory requirements. Each participant will be assigned a unique study identification number, and all study data, including clinical, behavioral, and biological data, will be de-identified using this code. No directly identifiable personal information will be used in analyses or stored with research data.

Hard copies of consent forms and any paper records will be stored in locked filing cabinets within a secure, access-restricted research facility. Electronic data will be stored on password-protected, encrypted institutional servers with access limited to authorized study personnel only.

Biological specimens (blood and stool) will be labeled using coded identifiers and stored separately from identifiable information. The linkage key between participant identities and study codes will be stored in a separate secure file accessible only to the principal investigator or designated authorized personnel.

All data transfers, analyses, and reporting will use de-identified datasets to ensure participant confidentiality is maintained throughout and after the study.

### Access to Data

Access to identifiable participant information will be restricted to trained research personnel involved in participant recruitment and in-person data collection. Each participant will be assigned a unique study identification code, and all datasets used for analysis will be de-identified.

De-identified data will be accessible to study investigators, statisticians, and bioinformaticians for data analysis. No directly identifiable information will be included in analytical datasets.

The key document between participant identifiers and study codes will be stored separately in a secure, access-restricted file maintained only by the principal investigator and authorized study personnel.

### Ancillary and Post-Trial Care

Given the low-risk nature of the intervention (supervised moderate-intensity aerobic exercise in a controlled laboratory setting), no specific ancillary or post-trial care is anticipated. The risks associated with participation are minimal and primarily related to transient exercise-related discomfort or venipuncture (e.g., soreness or mild bruising).

Participants will be advised to seek medical attention from their primary healthcare provider if they experience any adverse symptoms or concerns following study participation, and to share them as soon as possible with the study personnel. Any adverse events occurring during study procedures will be documented and reported to the Institutional Review Board in accordance with institutional policies.

### Participant Compensation

Participants will receive financial compensation for their time and participation in the study. Compensation will be provided regardless of study completion and will be prorated based on the number of completed study visits. Participants will be informed of the compensation structure during the informed consent process.

### Dissemination Policy

Study findings will be disseminated through presentations at scientific conferences and publication in peer-reviewed journals. Authorship will be determined in accordance with standard academic contribution guidelines.

De-identified microbiome sequencing data will be deposited in publicly accessible repositories (e.g., NCBI Sequence Read Archive or equivalent), and de-identified metabolomics data will be shared in appropriate public databases (e.g., MetaboLights or Metabolomics Workbench), in accordance with participant consent and institutional data sharing policies. All shared datasets will exclude any directly identifiable participant information.

## Discussion

The present manuscript describes a single-arm experimental pilot study investigating the acute effects of a single exercise bout on gut microbiome composition, function, and the gut-derived metabolome in premenopausal young adult women. George Washington University’s exercise and nutrition sciences physiology lab and genomics core set the environment for a successful delivery of the described protocol. The integration of an omics approach using novel methodologies such as BCGs to identify gut microbiome functionality, coupled with stool and plasma metabolome analyses, is unique and will provide insights into the acute response to an exercise bout. Obesity heterogeneity is a complex concept; investigating the gut microbiome and metabolomic contributors could enable future early identification of potential transitions from healthy to unhealthy states, as well as of responses to lifestyle interventions. To our knowledge, this is the first pilot study exploring the acute response to a bout of exercise across different obesity phenotypes in young-adult women.

Several challenges are anticipated. Recruitment of participants might be challenging, particularly for those participants who are sedentary, and due to the absence of any medication, considering ∼50% adults in the US have used a prescribed medication in the last 30 minutes who are prescribed one or more drugs in the US. To address this anticipated challenge, the protocol incorporates diverse recruitment strategies, provides detailed information before the baseline visit, and offers financial incentives for participation. Variability in the diet may also introduce heterogeneity to the gut microbiome composition and function. While we are aware that a feeding-controlled study would be ideal, we did not want to alter their habitual dietary intake, which would add additional changes to the gut microbiome. We introduced a food record form throughout the study, rather than a 24-hour dietary recall the day before stool collection, to capture their diet around the time of stool sample collection. An additional limitation is that pre-stratified phenotypes are based on self-reported data. We are aware of this limitation, but given the preliminary, hypothesis-generating nature of this pilot study, we will consider it in future larger studies. For example, having a 7-day running period to confirm their habitual physical activity would greatly benefit our future trial.

### Expected impact

We propose that an acute 30-minute aerobic exercise bout at 60-70% of HRR is likely to affect the relative abundance of SCFA-producing bacteria, potentially activating distinct metabolic pathways within their genomes, and contributing to increased SCFA in the gut, thereby releasing SCFAs into the bloodstream. Furthermore, the magnitude of change in gut microbiome and metabolome may be partially explained by the obesity phenotype. This research could contribute meaningfully to the field that almost 1 in 2 adults in the US has obesity, especially women, who face the highest prevalence of severe obesity.^1,2^ Obesity is a risk factor for several chronic diseases, including cardiovascular disease, diabetes, and cancer, and the associated healthcare burden is over $200 billion annually.^3^ By improving our understanding of the biological mechanisms underlying variability in exercise responses, this work may help inform more personalized strategies to improve cardiometabolic health. The roles of gut microbiome functional activity and gut-synthesized metabolites in tailoring lifestyle interventions for obesity and its heterogeneity remain largely unexplored yet are critical for developing personalized treatments. In particular, the utilization of gut-synthesized metabolites as potential early markers of a detrimental shift from a healthy obesity phenotype to a suboptimal one. Furthermore, using markers to understand individual responses to exercise could help us further improve exercise prescription for cardiometabolic health.

## Trial Status

This study is currently recruiting participants. The protocol is registered on ClinicalTrials.gov (NCT06691100). Recruitment is ongoing, and data collection has commenced.

## Supporting information

PARQ-questionnaire

PSQI-questionnaire

WALI-questionnaire

## Data Availability

No datasets were generated or analyzed during the preparation of this study protocol. Data generated during the course of the study will be available upon reasonable request, subject to ethical approval and institutional regulations governing the sharing of clinical research data. De-identified microbiome sequence data and metabolomic datasets generated in this study will be deposited in publicly available scientific repositories in accordance with journal and funding requirements. All shared data will be fully de-identified to protect participant confidentiality.

## List of Abbreviations Declarations

### Consent for publication

The consent form includes a section that states that the collected data will be used in an aggregated form, never as an individual data point, for research dissemination purposes. No images or videos of the participants are obtained.

### Authors contributions

Author contributions reflect each author’s role in the conceptualization, design, and planned implementation of this pilot study protocol. All authors participated in drafting and critical revision of the manuscript. All authors read and approved the final manuscript.

CPOS: Conceptualization, methodology, investigation, writing- original draft

DK: methodology, writing and editing

AR: writing and editing

KC: methodology, writing and editing

## Acknowledgements

We extend our sincere gratitude to the participants who have already finalized the study for their interest, time, and effort to contribute to science.

We are grateful to the research team in the FITGut lab, including Allison Chetta, Jordyn Walters, Andrea Osorio, Clara De Torres Enseñat, Gianna Filippou, Shery Kamel, and Karla Cordero-Ortega. Their expertise and time were fundamental in ensuring the ongoing development of the trial.

We want to thank the laboratory personnel at the GW Genomics Core (RRID: 027546), Jack Villani and Kamwing Jair, for their unconditional support for the preparation, processing, and sequencing of the biological samples.

