## Supplementary material for "Acute Exercise-Induced Changes in Gut Microbiome Composition, Function, and Gut-Derived Stool and Plasma Metabolome Across Obesity Phenotypes in Young-Adult Women: A Pilot Study Protocol": PARQ-questionnaire

### PAR-Q

Date ______________________

**The health benefits of regular physical activity are clear; more people should engage in physical activity every
day of the week. Participating in physical activity is very safe for MOST people. This questionnaire will tell you
whether you must seek further advice from your doctor OR a qualified exercise professional before becoming
more physically active.**

General Health Questions

Please, read the 7 questions below carefully and answer each one honestly: check YES or NO

Has your doctor ever said that you have a heart condition OR high blood pressure?
◯ Yes
◯ No

Do you feel pain in your chest at rest, during your daily living activities, OR when you do physical activity?
◯ Yes
◯ No

Do you lose balance because of dizziness OR have you lost consciousness in the last 12 months? (Please, answer NO if your dizziness was associated with over-breathing (including during vigorous exercise)
◯ Yes
◯ No

Have you ever been diagnosed with another chronic medical condition (other than heart disease or high blood pressure?
◯ Yes
◯ No

(If yes, please list the condition(s) here) ______________________

Are you currently taking prescribed medication for a chronic medical condition?
◯ Yes
◯ No

Do you currently have (or have had within the past 12 months) a bone, joint, or soft tissue (ligament, muscle, or tendon) problem that could be made worse by becoming more physically active? Please answer NO if you have a problem in the past, but it does not limit your current ability to be physically active
◯ Yes
◯ No

If yes, please list the condition(s) here. ______________________

Has your doctor ever said that you should only do medically supervised physical activity?
◯ Yes
◯ No

**If you answered No to all of the questions above, you are cleared for physical activity.**

I, the undersigned, have read and fully understood this questionnaire to my satisfaction and have completed it accurately. I acknowledge that this physical activity clearance is valid for up to 12 months from the date of completion and becomes invalid if my condition changes. I also acknowledge that the community/fitness center may retain a copy of this form for its records. In such instances, the center will maintain the confidentiality of the information in compliance with applicable laws.

Printed Name ______________________________

Signature of the participant ______________________________

Signature of the team ______________________________

Date ______________________________
