## Supplementary material for "Acute Exercise-Induced Changes in Gut Microbiome Composition, Function, and Gut-Derived Stool and Plasma Metabolome Across Obesity Phenotypes in Young-Adult Women: A Pilot Study Protocol": PSQI-questionnaire

### Pittsburg Sleep Quality Index Questions

1) Date ______________________

2) During the past month, what time have you usually gone to bed at night? ______________________

3) During the past month, how long (in minutes) has it usually taken you to fall asleep each night? (i.e., 45 minutes) ______________________

4) During the past month, what time have you usually gotten up in the morning? ______________________

5) During the past month, how many hours of actual sleep did you get at night? (This may be different than the number of hours you spent in bed) ______________________

6) During the past month, how often have you had trouble sleeping because you cannot get to sleep within 30 minutes?
- ◯ Not during the past month
- ◯ Less than once a week
- ◯ Once or twice a week
- ◯ Three or more times a week

7) During the past month, how often have you had trouble sleeping because you wake up in the middle of the night or early morning?
- ◯ Not during the past month
- ◯ Less than once a week
- ◯ Once or twice a week
- ◯ Three or more times a week

8) During the past month, how often have you had trouble sleeping because you have to get up to use the bathroom?
- ◯ Not during the past month
- ◯ Less than once a week
- ◯ Once or twice a week
- ◯ Three or more times a week

9) During the past month, how often have you had trouble sleeping because you cannot breathe comfortably?
- ◯ Not during the past month
- ◯ Less than once a week
- ◯ Once or twice a week
- ◯ Three or more times a week

10) During the past month, how often have you had trouble sleeping because you cough or snore loudly?
- ◯ Not during the past month
- ◯ Less than once a week
- ◯ Once or twice a week
- ◯ Three or more times a week

11) During the past month, how often have you had trouble sleeping because you feel too cold?
- ◯ Not during the past month
- ◯ Less than once a week
- ◯ Once or twice a week
- ◯ Three or more times a week

12) During the past month, how often have you had trouble sleeping because you feel too hot?
- ◯ Not during the past month
- ◯ Less than once a week
- ◯ Once or twice a week
- ◯ Three or more times a week

13) During the past month, how often have you had trouble sleeping because you have bad dreams?
- ◯ Not during the past month
- ◯ Less than once a week
- ◯ Once or twice a week
- ◯ Three or more times a week

14) In the past month, how often have you had trouble sleeping because you have pain?
- ◯ Not during the past month
- ◯ Less than once a week
- ◯ Once or twice a week
- ◯ Three or more times a week

15) If applicable, please describe other reasons why you have trouble sleeping.

__________________________________

16) If applicable, please list how often you have trouble sleeping for these other reasons.
- ◯ Not during the past month
- ◯ Less than once a week
- ◯ Once or twice a week
- ◯ Three or more times a week

17) During the past month, how would you rate your sleep quality overall?
- ◯ Very good
- ◯ Fairly good
- ◯ Fairly bad
- ◯ Very bad

18) During the past month, how often have you taken medicine to help you sleep (prescribed or "over the counter") ?
- ◯ Not during the past month
- ◯ Less than once a week
- ◯ Once or twice a week
- ◯ Three or more times a week

19) During the past month, how often have you had trouble staying awake while driving, eating meals, or engaging in social activity?
- ◯ Not during the past month
- ◯ Less than once a week
- ◯ Once or twice a week
- ◯ Three or more times a week

20) During the past month, how much of a problem has it been for you to keep up enough enthusiasm to get things done?
- ◯ No problem at all
- ◯ Only a very slight problem
- ◯ Somewhat of a problem
- ◯ A very big problem

21) Do you have a bed partner or room mate?
- ◯ No bed partner or room mate
- ◯ Partner/room mate in other room
- ◯ Partner in the same room, but not same bed
- ◯ Partner in same bed

22) If you have a room mate or bed partner, ask him/her how often in the past month you have had loud snoring.
- ◯ Not during the past month
- ◯ Less than once a week
- ◯ Once or twice a week
- ◯ Three or more times a week

23) If you have a room mate or bed partner, ask him/her how often in the past month you have had long pauses between breaths of sleep.
- ◯ Not during the past month
- ◯ Less than once a week
- ◯ Once or twice a week
- ◯ Three or more times a week

24) If you have a room mate or bed partner, ask him/her how often in the past month you have had legs twitching or jerking while you sleep.
- ◯ Not during the past month
- ◯ Less than once a week
- ◯ Once or twice a week
- ◯ Three or more times a week
