## Supplementary material for "Acute Exercise-Induced Changes in Gut Microbiome Composition, Function, and Gut-Derived Stool and Plasma Metabolome Across Obesity Phenotypes in Young-Adult Women: A Pilot Study Protocol": WALI-questionnaire

### Lifestyle Questionnaire

Date ______________________

Do you follow a prescribed exercise training?
◯ Yes
◯ No

What exercise/sports training do you follow? ______________________

How many hours do you exercise/train per week? ______________________

How long does your exercise training per session last in minutes? ______________________

Do you use any electronic tracking method to track your training load?
◯ Yes
◯ No

If yes, please indicate which one ______________________

Are you taking any dietic supplements (e.g., fiber, multivitamins, probiotics)?
◯ Yes
◯ No

If yes, please indicate which one. ______________________

Do you follow any particular diet?
☐ Vegan
☐ Vegetarian (Including dairy and eggs)
☐ Pescatarian (Including dairy, eggs, and fish)
☐ Ketogenic Diet
☐ Omnivore
☐ Paleo Diet
☐ Other
☐ None

The following questions will address questions about your weight history, eating habits, eating patterns, and psychological factors

#### Weight history

Have you ever tried to lose weight on your own or with a professional?
◯ Yes
◯ No

How much weight did you lose?

______ lbs.

At what weight did you start to diet during this time?

______ lbs.

Which statement best describes you? "During the past 6 months, my weight has..."

- ◯ decreased more than 10 lbs. or more
- ◯ decreased by 5 to 10 lbs.
- ◯ been relatively stable
- ◯ increased by 5 to 10 lbs.
- ◯ increased by more than 10 lbs. or more

---

Have you ever been told at any given point in your life that your weight was not healthy for your age and height?

- ◯ Yes
- ◯ No

---

At what age were you first overweight by 10 lbs or more?

__________________________________

(The age when you noticed you were above the ideal or healthy weight for your heigh and body type)

---

How do you remember that you were overweight at the time? (e.g., pictures, clothing size, others telling you)

__________________________________________

---

What has been your highest weight after the age of 21?

lbs. yrs old.

______ ______

---

What has been your lowest weight (not due to illness) after the age of 21, which you have maintained for at least 1 year?

lbs. yrs. old maintained for ______ yrs.

______ ______ ______

---

Was this weight reached after a weight loss effort?

- ◯ Yes
- ◯ No

---

What was your weight:

6 months ago? 1 year ago? 2 years ago?

______ lbs. ______ lbs. ______ lbs.

---

For each time period shown, please list your maximum weight. If you cannot remember what your maximum weight way, make your best guess and mark "G" (for guess) next to your answer. In addition please note any events related to your gaining weight during this period. For ages 16 and beyond, please identify the figure, from the attached image below, that most resembles your figure at that time. Record your number of the figure.

Age Maximum Weight Figure # Events Related to Weight Gain

5-10 ______

11-15 ______

16-20 ______ ______ ______

21-25 ______ ______ ______

26-30 ______ ______ ______

31-35 ______ ______ ______

36-40 ______ ______ ______

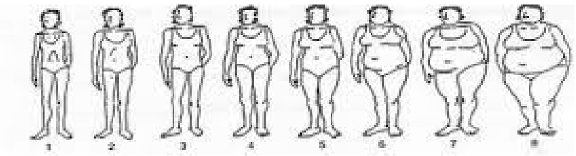

#### Family Weight History

Please indicate the average height and weight of your biological mother and father during their middle-age years.

Parent Height (ft. + in.) Weight (lbs.) Current age or year of death
Mother ______ ______ ______
Father ______ ______ ______

Please indicate the height and weight of the following members in your immediate family. Indicate any half-brothers or half-sisters.

Family Member Height (ft. + in.) Weight (lbs.) Current age of year of death
Spouse/Significant Other ______ ______ ______
Older brother ______ ______ ______
2nd oldest brother ______ ______ ______
3rd oldest brother ______ ______ ______
Oldest sister ______ ______ ______
2nd oldest sister ______ ______ ______
3rd oldest sister ______ ______ ______

#### Weight Loss History

Please record your major weight loss efforts (i.e., diet, exercise, moderation, etc.) that resulted in a weight loss of 10 pounds or more. Take time to think over your previous efforts, starting with the first one, whether in childhood or adulthood. You may have difficulty remembering this information at first, but most people can if they take their time. Start with our first weight loss effort and proceed in order until you reach your most recent one.

If you have never engage in any weight loss program, please jump to the next section, WEIGHT, PREGNANCY, AND MENSTRUAL CYCLE

Age at time of effort Weight at the start of effort # of lbs. lost Method used to lose weight
______ ______ ______ ______
______ ______ ______ ______
______ ______ ______ ______
______ ______ ______ ______
______ ______ ______ ______

Please pick a number from 1 to 10 to indicate below how accurate you think you were in remembering and recording your weight loss history. Pick any number from 1 to 10.

1 = not at all accurate
10 = completely accurate

In the past year, how many times have you started a weight loss program on your own that lasted for more than three days? __________________________________

In the past year, how many times have you started a weight loss program that lasted for 3 days or less? __________________________________

Have you ever experienced any significant physical or emotional symptoms while attempting to lose weight or after losing weight?
◯ Yes
◯ No

If yes, please describe your symptoms, how long they have lasted and the type of professional help sought, if any.

Problem Year Duration (wks.) Type of Professional Help
______ ______ ______ ______
______ ______ ______ ______
______ ______ ______ ______

#### Weight, Pregnancy, and Menstrual Cycle

Have you ever borne children?
◯ Yes
◯ No

What was the weight at the start of your pregnancy?

______ lbs.

What was your weight at delivery?

______ lbs.

What was your lowest at delivery?

______ lbs.

What was your weight at the start of your second delivery?

______ lbs

What was your weight at delivery?

______ lbs.

What was your lowest weight after delivery?

______ lbs.

What was your weight at the start of your third pregnancy?

______ lbs.

What was your weight at delivery?

______ lbs.

What was your lowest weight after delivery?

______ lbs.

Do you experience a regular menstrual cycle?
◯ Yes
◯ No

If yes, please describe your eating around the time of your menstruation.
◯ Eat much less
◯ Eat less
◯ No change
◯ Eat more
◯ Eat much more

Do you crave particular foods around the time of your menstruation?
◯ Yes
◯ No

If yes, which foods do you crave?

__________________________________

#### Eatings Habits

Please indicate the degree to which you believe each of the following behaviors causes you to gain weight. To answer these questions, please use the 5-point scale below. Pick the number that best describes how much the behavior contributes to your increased weight gain.

- does not contribute at all
- contributes a small amount
- contributes a moderate amount
- contributes a large amount
- contributes the greatest amount

Eating with family/friends ______
Eating when socializing/celebrating ______
Eating at business functions ______
Eating when happy ______
Eating in response to the sight or smell of food ______
Eating because of the good taste of food ______
Eating because I can't stop once I've begun ______
Overeating at dinner ______
Eating too much food ______
Continuing to eat because I don't feel full after a meal ______
Eating because I crave certain foods ______
Eating because I feel physically full ______
Eating while cooking/preparing food ______
Eating when stressed ______
Eating when depressed/upset ______
Eating when angry ______
Eating when anxious ______
Eating when alone ______
Eating when bored ______
Eating when tired ______
Overeating at lunch ______
Overeating at breakfast ______
Snacking after dinner ______
Snacking between meals ______

Please indicate any other factors that contribute to a moderate amount or more to your weight gain.

__________________________________

How many days a week do you eat the following meals? Write the number of days in the space and the usual time of each meal.

Meals Days a Week Time Snacks Days a Week Time
Breakfast ______ ______ Morning Snack ______ ______
Lunch ______ ______ Afternoon Snack ______ ______
Dinner ______ ______ Evening Snack ______ ______

Who prepares meals at your home?

__________________________________

Who does the food shopping?

__________________________________

Please list your five favorite foods.

__________________________________

Do you have any food allergies?

◯ Yes
◯ No

If yes, please specify the food and the allergic reactions.

__________________________________________

Please specify the amount (in cps, 8 oz) of the following fluids you typically consume a day.

______ skim milk ______ low-fat milk ______ whole milk ______ seltzer water ______ beer
______ fruit juice ______ diet soda ______ tea ______ coffee ______ other
______ water ______ regular soda ______ wine ______ hard liquor

During a typical week, how many meals do you eat at a fast food restaurant (including drive-thru and convenience stores) ?

Breakfast (meals a week) ______
Lunch (meals a week) ______
Dinner (meals a week) ______

During a typical week, how many meals do you eat at a traditional restaurant, coffee shop, cafeteria, or similar establishment?

Breakfast (meals a week) ______
Lunch (meals a week) ______
Dinner (meals a week) ______

How many times a week do you typically eat out with others (including family)?

__________________________________

#### Eating Patterns I

During the past 6 months, did you often eat an unusually large amount if food within a two hour period (an amount that most people would agree is unusually large)?

◯ Yes
◯ No

During the times when you ate an unusually large amount of food, did you often feel you could not stop eating or control what or how much you were eating?

◯ Yes
◯ No

During the past 6 months, how often, on average, did you have times when you ate unusually large amounts of food and felt that your eating was out of control? (There may have been some weeks when it was not present so just average those in.)

◯ Less than one day a week
◯ One day a week
◯ Two or three days a week
◯ Four or five days a week
◯ Nearly every day

Did you usually have any of the following experiences during these occasions?

Eating much more rapidly than usual?

◯ Yes
◯ No

Eating until you felt uncomfortably full?

◯ Yes
◯ No

Eating large amounts of food when you didn't feel physically hungry?

◯ Yes
◯ No

Eating alone because you were embarrassed by how much you were eating?

◯ Yes
◯ No

Feeling disgusted with yourself, depressed or feeling very guilty after overeating?

◯ Yes
◯ No

Eating large amounts of food throughout the day with no planned mealtimes?

◯ Yes
◯ No

Think about a typical time when you ate this way (that is, large amounts of food and feeling that your eating was out of control.) What time of the day did the episode start?

◯ Morning (8 AM to 12 Noon)
◯ Early afternoon (12 Noon to 4 PM)
◯ Late afternoon (4 PM to 7 PM)
◯ Evening (7 PM to 10 PM)
◯ Night (After 10 PM)

Approximately how long did this episode of eating last? From the time you started to eat until when you stopped and did not eat again for at least two hours?

Hours Minutes

______ ______

As best as you can remember, please list everything you might have eaten or drunk during that episode. If you ate for more than two hours, describe the food and liquids you consumed the most. Be specific and include amounts and brands (when possible). Estimate as best as you can.

For example, 7 ounces of Ruffles potato chips; 1 cup of Breyer's chocolate ice cream with 2 teaspoons of hot fudge; two 8-ounce glasses of Coca-Cola; and 1 and a half ham and cheese sandwiches with mustard.

| Food | Amount | Brand (if possible) |
|---|---|---|
| ______ | ______ | ______ |
| ______ | ______ | ______ |
| ______ | ______ | ______ |
| ______ | ______ | ______ |
| ______ | ______ | ______ |
| ______ | ______ | ______ |
| ______ | ______ | ______ |

At the time this episode started, how long had it been since you had previously finished eating a meal or snack?

______ hours ______ minutes

In general, during the past 6 months, how upset were you by overeating episodes in which you ate unusually large amounts of food?

◯ Not at all
◯ Slightly
◯ Moderately
◯ Greatly
◯ Extremely

In general, during the past 6 months how upset were you by the feeling that you could not stop eating or could not control what or how you were eating?

◯ Not at all
◯ Slightly
◯ Moderately
◯ Greatly
◯ Extremely

In general, during the past 6 months, how important has your weight or shape been in how you feel about or evaluate yourself as a person-compared to other aspects of your life (i.e. how do you work as a parent, or how you get along with other people)? Weight and shape...

◯ were not very important
◯ played a part in how I feel about myself
◯ were among the main things that affected how I felt about myself
◯ were the most important things that affected how I felt about myself

| | |
|---|---|
| During the past 3 months, did you ever make yourself vomit in order to avoid gaining weight after binge eating? | ◯ Yes<br>◯ No |
| If yes, how often, on average, was that? | ◯ Less than once a week<br>◯ Once a week<br>◯ Two or three times a week<br>◯ Four or five times a week<br>◯ More than five times a week |
| During the past 3 months, did you ever take more than twice the recommended dose of laxatives in order to avoid gaining weight after binge eating? | ◯ Yes<br>◯ No |
| If yes, how often, on average, was that? | ◯ Less than once a week<br>◯ Once a week<br>◯ Two or three times a week<br>◯ Four or five times a week<br>◯ More than five times a week |
| During the past 3 months, did you ever take more than twice the recommended dose of diuretics (water pills) in order to avoid gaining weight after binge eating? | ◯ Yes<br>◯ No |
| If yes, how often, on average, was that? | ◯ Less than once a week<br>◯ Once a week<br>◯ Two or three times a week<br>◯ Four or five times a week<br>◯ More than five times a week |
| During the past 3 months, did you ever fast (not eat anything at all for at least 24 hours) in order to avoid gaining weight after binge eating | ◯ Yes<br>◯ No |
| If yes, how often, on average, was that? | ◯ Less than once a week<br>◯ Once a week<br>◯ Two or three times a week<br>◯ Four or five times a week<br>◯ More than five times a week |
| During the past 3 months, did you ever exercise for more than one hour specifically in order to avoid gaining weight after eating? | ◯ Yes<br>◯ No |
| If yes, how often, on average, was that? | ◯ Less than one day a week<br>◯ One day a week<br>◯ Two or three days a week<br>◯ Four or five days a week<br>◯ Nearly every day |
| During the past 3 months, did you ever take more than twice the recommended dosage of a diet pill in order to avoid gaining weight after binge eating? | ◯ Yes<br>◯ No |
| If yes, how often, on average, was that? | ◯ Less than once a week<br>◯ Once a week<br>◯ Two or three times a week<br>◯ Four or five times a week<br>◯ More than five times a week |

#### Eating Patterns II

How hungry are you usually in the morning?

- 0 - Not at all
- 1 - A little
- 2 - Somewhat
- 3 - Moderately
- 4 - Very

When do you usually eat for the first time?

- 0 - Before 9 AM
- 1 - 9:01 to 12 PM
- 2 - 12:01 - 3PM
- 3 - 3:01 to 6 PM
- 4 - 6:01 PM or later

Do you have cravings or urges to eat snacks after supper, but before bedtime?

- 0 - Not at all
- 1 - A little
- 2 - Somewhat
- 3 - Moderately
- 4 - Very

How much control do you have over your eating between supper and bedtime?

- 0 - Not at all
- 1 - A little
- 2 - Somewhat
- 3 - Moderately
- 4 - Very

How much of your daily food intake do you consume after suppertime?

- 0 - 0% (none)
- 1 - 1-25% (up to a quarter)
- 2 - 26-50% (about half)
- 3 - 51-75% (more than half)
- 4 - 76 - 100% (almost all)

Are you currently feeling blue or down in the dumps?

- 0 - Not at all
- 1 - A little
- 2 - Somewhat
- 3 - Moderately
- 4 - Very

When you are feeling blue, is your mood lower in the:

- 0 - Early Morning
- 1 - Late Morning
- 2 - Afternoon
- 3 - Early Evening
- 4 - Late Evening/Night

How often do you have trouble getting to sleep?

- 0 - Never
- 1 - Sometimes
- 2 - About half of the time
- 3 - Usually
- 4 - Always

Other than only to use the bathroom, how often do you get up at least once in the middle of the night?

- 0 - Never
- 1 - Less than once a week
- 2 - About once a week
- 3 - More than once a week
- 4 - Every night

Do you have cravings or urges to eat snacks when you wake up?

- 0 - Not at all
- 1 - A little
- 2 - Somewhat
- 3 - Very much so
- 4 - Extremely

Do you need to eat in order to get back to sleep when you awake at night?

- ◯ 0 - Not at all
- ◯ 1 - A little
- ◯ 2 - Somewhat
- ◯ 3 - Very much so
- ◯ 4 - Extremely

When you get up in the middle of the night, how often do you snack?

- ◯ 0 - Never
- ◯ 1 - Sometimes
- ◯ 2 - About half the time
- ◯ 3 - Usually
- ◯ 4 - Always

When you snack in the middle of the night, how aware are you of your eating?

- ◯ 0 - Not at all
- ◯ 1 - A little
- ◯ 2 - Somewhat
- ◯ 3 - Very much so
- ◯ 4 - Completely

How much control do you have over your eating while you are up at night?

- ◯ 0 - None at all
- ◯ 1 - A little
- ◯ 2 - Same
- ◯ 3 - Very much
- ◯ 4 - Complete

How long have your difficulties with night eating has been going on?

______ months ______ years

#### Physical Activity

To what extent do you enjoy physical activity?

- ◯ not at all
- ◯ slightly
- ◯ moderately
- ◯ greatly

Do you have any physical problems that limit your physical activity?

- ◯ Yes
- ◯ No

If yes, please describe.

__________________________________________

Please check the types of physical activity that you enjoy. Only check those that you have participated in during the last year.

- ☐ walking outside
- ☐ walking (indoors, including treadmill)
- ☐ jogging
- ☐ running
- ☐ biking outside
- ☐ biking (stationary)
- ☐ aerobic class
- ☐ tennis/racket sports
- ☐ swimming
- ☐ basketball
- ☐ golf
- ☐ dancing
- ☐ strength training
- ☐ other

For your preferred activity, how many times have you participated in this activity in the past 6 months?

______ times

How many hours of TV do you watch on an average weekday?

______ hours

How many hours of TV do you watch on an average weekend day?

______ hours

Approximately how many city blocks or the equivalent do you regularly walk each day? (12 blocks = 1 mile)

______ blocks

How many flights of stairs do you climb each day? (1 flight = 10 steps)

______ flights

Please describe you daily lifestyle activity (i.e., how active you are) by picking any number from 1 to 1 in which 1 = very sedentary and 10 = very active. Your number is ______________________________

#### Self-Perceptions

How satisfied are you with your current weight?

◯ very satisfied
◯ moderately satisfied
◯ slightly satisfied
◯ neutral
◯ slightly dissatisfied
◯ moderately dissatisfied
◯ very dissatisfied

How satisfied are you with your current shape? (i.e., figure or physique)?

◯ very satisfied
◯ moderately satisfied
◯ slightly satisfied
◯ neutral
◯ slightly dissatisfied
◯ moderately dissatisfied
◯ very dissatisfied

How satisfied are you with your current overall appearance?

◯ very satisfied
◯ moderately satisfied
◯ slightly satisfied
◯ neutral
◯ slightly dissatisfied
◯ moderately dissatisfied
◯ very dissatisfied

Pick the one sentence that best describes your overall feelings about yourself. "In general, I am..."

◯ very happy with who I am
◯ happy with who I am
◯ okay with who I am, but have some mixed feelings
◯ unhappy with who I am
◯ very unhappy with who I am

"As compared to most people, I think I have..."

◯ very good self-esteem
◯ good self-esteem
◯ average self-esteem
◯ poor self-esteem
◯ very poor self-esteem

Pick one sentence that best describes your feelings about the way you look the last time you lost a lot of weight. "I was..."

- ◯ very happy with the way I looked
- ◯ happy with the way I looked
- ◯ okay with the way I looked, but with some mixed feelings
- ◯ unhappy with the way I looked
- ◯ very unhappy with the way I looked

#### Psychological Factors

Have you ever had any problems at any time with depression, anxiety, or any other emotions that disrupted your normal functioning?

- ◯ Yes
- ◯ No

Have you ever sought professional help for emotional problems?

- ◯ Yes
- ◯ No

If yes, specify below.

Problem Year Duration (wks.) Type of Professional Help

______ ______ ______ ______
______ ______ ______ ______
______ ______ ______ ______
______ ______ ______ ______

During the past month, have you felt depressed, sad, or blue much of the time?

- ◯ Yes
- ◯ No

During the past month, have you often felt hopeless about the future?

- ◯ Yes
- ◯ No

During the past month, have you had little interest or pleasure in doing things?

- ◯ Yes
- ◯ No

#### Timing

Please indicate if you are currently experiencing any greater than usual stress in your life related to the following events.

Work

- ◯ Yes
- ◯ No

Health

- ◯ Yes
- ◯ No

Relationship with spouse/significant other

- ◯ Yes
- ◯ No

Activities related to your children

- ◯ Yes
- ◯ No

Activities related to your parents

- ◯ Yes
- ◯ No

Legal/financial trouble

- ◯ Yes
- ◯ No

| | |
|---|---|
| School | ◯ Yes<br>◯ No |
| Moving | ◯ Yes<br>◯ No |
| Other (please explain) | ______________________________________ |
| Are you planning any major life changes (i.e., new job, moving, relationship, etc.) during the next 6 months? | ◯ Yes<br>◯ No |
| If yes, please briefly explain: | ______________________________________ |
| How stressful has your life been during the past 6 months? | ◯ much less stressful than usual<br>◯ less stressful than usual<br>◯ average level of stress<br>◯ more stressful than usual<br>◯ much more stressful than usual |
